# A Quantitative Capability and Needs Assessment Across 12 African Health and Demographic Surveillance Systems under INSPIRE

**DOI:** 10.64898/2025.11.30.25341326

**Authors:** Agnes Kiragga, Samuel Iddi, Ivan Busulwa, Rachel Odhiambo, Henry Owoko Odero, Daniel Maina, INSPIRE, Damazo Kadengye

## Abstract

**Background:** Health and demographic surveillance systems (HDSS) provide essential longitudinal population data in contexts where civil registration and administrative systems are incomplete. Despite their importance, HDSS data systems vary substantially in infrastructure, governance, and analytic capacity. As part of the Implementation Network for Sharing Population Information from Research Entities (INSPIRE) 2.0 program, we conducted a comprehensive capability and needs assessment across 12 African HDSS in 10 countries to document current data ecosystems, capability maturity, and training needs.

**Methods:** A quantitative needs assessment was conducted between April - June, 2025 using a REDCap-based survey completed by HDSS technical staff, including site leads, data managers, and data analysts. The survey captured standardized metrics across four data management domains, namely, site characteristics, data ecosystem, capability maturity, and workforce training needs. Data were analyzed descriptively in R, with cross-site comparisons used to identify patterns, gaps, and priority areas for investment.

**Results:** Across the 12 sites, the assessment revealed strong adoption of electronic data collection tools but persistent reliance on hybrid paper-digital workflows at one-third of sites. Metadata standards, interoperability mechanisms, automation pipelines, and cloud infrastructures were inconsistently implemented. Capability maturity varied widely across data management, governance, platforms, and user culture. Training needs were substantial across all seven domains, especially in research and methodology, technology and infrastructure, and data analytics. The analysis identified systemic gaps in coding capacity, metadata standards, governance monitoring, ETL automation, cloud readiness, and advanced analytics.

**Conclusions:** HDSS platforms in Africa maintain robust operations but require significant capacity strengthening in governance, automation, metadata management, and analytics to achieve higher data capability maturity. Workforce development must be prioritized through structured, bilingual training programs aligned with INSPIRE 2.0. These findings provide a critical baseline for designing targeted interventions and guiding harmonization of HDSS data systems in Africa.

## Introduction

Health and Demographic Surveillance Systems (HDSS) play a critical role in generating longitudinal population-level data in low- and middle-income countries, particularly where civil registration and vital statistics systems remain incomplete or fragmented (De Savigny et al., 2017). These systems provide repeated household-based enumeration and monitoring of births, deaths, migration, and health indicators. Over the past decade, HDSS sites have become important pillars of population data science in sub-Saharan Africa, supporting national statistical agencies, ministries of health, and global research consortia (Sankoh & Byass, 2012). With shifts in funding, including reduced U.S. support for demographic and health surveys in Africa, HDSS sites are emerging as key alternative data sources for national health policy (Khaki et al., 2025). Despite these strengths, HDSS data systems vary substantially in maturity across the African continent. Differences in infrastructure, data governance, metadata standards, technical capacity, interoperability, and workflow automation constrain harmonization and limit the scalability of analytic processes (Hinga et al., 2021). Recent developments in cloud computing, AI-assisted data processing, and automated data quality systems present new opportunities for modernizing HDSS platforms (Jiang et al., 2017). However, the degree to which African HDSS sites are prepared to adopt these innovations remains unclear.

The Implementation Network for Sharing Population Information from Research Entities (INSPIRE) 2.0 seeks to strengthen HDSS data ecosystems across Africa by supporting modernization, workforce development, and the introduction of automated and AI-enhanced data management tools. To inform this effort, we conducted a comprehensive capability and needs assessment across 12 African HDSS sites in 10 countries. This manuscript presents the findings of that assessment, with a focus on site characteristics, data ecosystems, capability maturity, and workforce training requirements.

### Background

HDSS platforms have evolved over decades, often shaped by donor priorities, national research agendas, and historical investment patterns (Sankoh & Byass, 2012). This evolution has produced a landscape characterized by methodological consistency in field operations, but substantial heterogeneity in data systems, governance frameworks, analytics readiness, and IT infrastructure (Ramsay et al., 2024). Challenges identified in the literature and echoed in our assessment include fragmented metadata systems, non-standardized data quality frameworks, limited interoperability with national health information systems, inconsistent governance structures, and limited adoption of modern anonymization techniques such as differential privacy or synthetic data (Chepkirui et al., 2025; Musa et al., 2023; Ramsay et al., 2024; Sankoh & Byass, 2012). Many HDSS sites retain manual or semi-manual data processes and rely on spreadsheets for data management, limiting their ability to scale and integrate with broader national systems (Oni et al., 2016). The INSPIRE 2.0 assessment was designed to quantify these gaps, characterize site maturity levels, and guide strategic, evidence-based capacity strengthening across the network.

## Methods

### Study design and setting

We conducted a cross-sectional, multi-site capability and needs assessment across 12 Health and Demographic Surveillance System (HDSS) sites participating in the INSPIRE 2.0 network. The assessment was designed to characterize each site’s data ecosystem, data governance structures, technological architecture, data capability maturity, and training needs. A structured, multi-domain questionnaire was administered through REDCap between April and June 2025. The assessment tool was developed jointly by the African Population and Health Research Center (APHRC) and INSPIRE 2.0 partners and underwent iterative review two months prior to deployment to ensure content validity, clarity, and alignment with FAIR and OMOP global data management frameworks.

### Participants and Sampling

The target respondents were individuals with institutional knowledge of HDSS operations, including site leads, data managers, data analysts, research managers, and ICT officers. Each participating HDSS provided a minimum of two respondents: (1) a Site Lead responsible for organizational information and high-level data processes, and (2) a Data Manager responsible for technical, governance, and platform-specific domains. Additional respondents (e.g., statisticians, analysts, ICT officers) contributed to specialized sections such as data architecture, metadata management, and infrastructure needs.

### Quantitative data collection

A detailed semi-structured survey was administered through REDCap to HDSS site leads, data managers, statisticians, information technology (IT) officers, and analysts. The survey captured site demographic characteristics, data sources and collection tools, IT infrastructure, data governance and security practices, anonymization techniques, data sharing agreements, data quality assurance, workforce composition, and capability maturity indicators. Quantitative data were exported to R for descriptive analysis.

### Data collection tool

Data for this assessment were collected using a comprehensive, multi-domain instrument developed to capture both quantitative indicators and contextual qualitative information. The tool covered institutional characteristics, data ecosystem functions, capability maturity, training needs, infrastructure gaps, and governance-related attitudes. The first component gathered core site and research organizational characteristics, including HDSS history, population coverage, staffing patterns, funding sources, surveillance frequency, operational costs, and the structure of data management units. A second component examined the full data ecosystem across four domains, i.e., Data, Platforms, Governance, and Users, documenting data sources and collection tools, software used for cleaning and analytics, validation and quality processes, storage architectures, security practices, metadata systems, and workforce competencies.

A structured maturity assessment formed the third component, scoring each site across People & Culture, Data Activities, Business Processes, and Technology using a standardized 0-4 scale aligned with established maturity models. This was followed by a detailed training needs module, in which sites rated the level of support required across governance, data, platform, and user/culture competencies, including areas such as data protection, harmonization, cloud systems, machine learning, metadata management, and visualization.

HDSS infrastructure needs were assessed through a dedicated section that captured the availability and functionality of hardware, software, connectivity, storage capacity, backup systems, and analytic tools. Additional modules evaluated data-sharing trust and governance readiness, including willingness to share data, perceived risks, and confidence in partners. Altogether, these components provided a holistic view of HDSS data systems, workforce skills, governance practices, and infrastructure readiness across the network.

### Conceptual framework

Our methodology was guided by a data ecosystem framework that outlines the core components required for robust, modern population data systems. Drawing on global data ecosystem models, the framework assesses HDSS capacities across four domains: data governance, infrastructure and platforms, data activities and user access, and innovation readiness, including emerging tools such as automation and AI. This framework provided a structured approach for evaluating site strengths, identifying operational and governance gaps, and determining areas requiring targeted investment. It emphasizes alignment with ethical and regulatory standards, sustainable and scalable infrastructure, secure data management practices, and improved interoperability. It also highlights the importance of user-oriented processes such as data sharing, accessibility, and collaboration across sites. By applying this framework, we systematically assessed the digital and organizational readiness of each HDSS, enabling the identification of priority areas for capability strengthening and future modernization.

### Data management

Data were extracted from REDCap and exported into R (version 4.3) for cleaning, processing, and analysis. Validation checks included range checks, consistency checks across related fields, and cross-respondent harmonization within HDSS sites. Open-ended responses were coded using an inductive/deductive approach, and quantitative elements were summarized using descriptive statistics.

### Analytical framework

HDSS capability maturity was assessed across four domains: People and Culture, Data Activities, Business Processes, and Technology. Scores were standardized on a five-point scale using the Higher Education Statistics Agency (HESA) toolkit, reflecting five progressive stages of maturity from chaotic/reactive, through stable, to proactive/predictive.

## Results

### Site characteristics

The fourteen participating HDSS sites covered rural, peri-urban, and urban settings and collectively surveilled more than 1.3 million people. Sites varied markedly in age, with the oldest established in 1970 and the newest launched in 2024, indicating a wide range of establishment dates. However, most sites were established between the 1990s and 2000s. Most sites conduct annual or biannual data collection rounds. Costs per round ranged widely, with a median around USD 60,000. Staffing profiles also varied, with total staff per site ranging from five to more than two hundred comprising a mixture of permanent and temporary personnel. Each HDSS is typically run by a national research institute or university in combination with other local or international research organizations, highlighting strong partnerships between local centers and international collaborators. These characteristics contextualize the diversity of technical and operational capacity observed across the network.

#### Number of households and population

Over time, HDSS populations have expanded significantly due to natural population growth and, in some cases, geographic expansion. At inception, the HDSS sites collectively covered approximately 594,000 people (median =36,000 per site, ~7,500 households). However, their current combined population is about 1.3 million, with a median of ~95,000 people per site; several larger sites now exceed 150,000-200,000 individuals. On average, sites have more than doubled in size from roughly 50,000 to 108,000 residents under surveillance underscoring the expanding scope of HDSS over time. One site (Magu HDSS) did not report current population data, suggesting inactivity or missing information.

#### Urban and rural composition of HDSS

Most HDSS sites continue to operate primarily in rural settings: 11 of the 12 sites include rural populations, consistent with HDSS historical focus on rural health and demographic monitoring. Nevertheless, urban coverage is also substantial, with six sites including urban populations, either in informal settlements, formal areas, or both. Only one site (Ouagadougou HDSS in Burkina Faso) is exclusively urban. Two additional sites described their contexts as peri-urban or semi-urban, reflecting transitional areas between rural and urban zones. Overall, while HDSS systems were traditionally rural, many have expanded to include urban and mixed populations.

#### Population age structure

Age-disaggregated data from eight of the 12 HDSS sites show a predominantly young population. Children under five typically constitute 15-20% of residents, while adults aged 50 years and above account for about 5-10%. This pattern aligns with demographic profiles in many African countries, where high fertility and improving, but still limited, life expectancy produce a broad-based, youthful age pyramid. The demographic structure underscores the relevance of maternal, child, and adolescent health research within HDSS settings. Four sites did not report age breakdowns, but the youthful pattern is consistent across both rural and urban sites that provided data.

#### Funding and institutional support

HDSS sites rely on a diverse mix of funders, with sites typically supported by four institutions to six. Funding comes from major international donors, national governments, research grants, and academic partners. Common funders include the Wellcome Trust, the Bill & Melinda Gates Foundation (often through continent-wide initiatives such as CHAMPS), the U.S. National Institutes of Health, UK research bodies such as UKRI and the Medical Research Council, as well as Sida (Sweden) and IDRC (Canada). Local governments and health facilities often contribute in-kind resources. Collectively, this mix illustrates that HDSS sustainability depends on broad, multi-partner funding arrangements supporting both core operations and project-specific activities.

#### Cost per surveillance round

Reported costs for a single HDSS surveillance round vary widely across sites, reflecting differences in population size, geography, staffing, and operational models. Estimates range from approximately USD 10,000 to USD 800,000 per round. Most sites fall within the USD 20,000-100,000 range, with a median of USD 60,000, typical of medium-sized (~50,000 population) HDSS operations. A few sites report substantially higher costs, over USD 400,000 and up to USD 800,000, often corresponding to large urban settings or enhanced data collection activities such as biobanking or clinical modules. While most HDSS rounds operate below USD 100,000, cost variation is driven by factors such as population size, geographic spread, transport needs, staffing levels, and the complexity of data collection.

#### Human resources and infrastructure

HDSS implementation is highly labor-intensive, with staffing levels varying across sites. Total staff numbers range from as few as five personnel to nearly 200, with a median of about 30 staff per site. Larger operations, such as AL-SEHA in Egypt and Hararghe in Ethiopia, maintain sizeable teams of up to 200 staff, while smaller or newer sites operate with minimal staff. Most sites combine a core group of permanent employees with a larger cohort of temporary fieldworkers hired during surveillance rounds. Typically, 40-60% of staff are permanent, though reliance on temporary staff increases where funding is project-based. Data management staff constitute a substantial share of these teams, averaging around half of all technical personnel, reflecting the central role of data processing and curation in maintaining HDSS longitudinal databases. Data management teams show moderate gender balance, with women making up about 42% of staff across reporting sites. Most sites (10 of 12) have a dedicated data management unit, usually centralized, while only two operate without a formal department, relying instead on distributed or ad hoc arrangements.

Every site identified a designated data management lead, highlighting the central role of specialized personnel in managing HDSS longitudinal data.

#### Longitudinal data availability and utilization

Over half of HDSS sites maintain multi-year household and population records, with seven out of 12 sites providing annual household counts and mid-year population data for 2020-2024. These data generally allow tracking of demographic changes such as migration and household formation and reflect the steady HDSS growth over time. Sites with both urban and rural components often report annual figures disaggregated by setting, with rural households and populations typically dominating the data sets. Sex-disaggregated population data, available for many sites, show roughly equal numbers of males and females, with a slight female majority in some cases. A subset of sites also supplied longitudinal age-group distributions, demonstrating capacity to monitor shifts in population structure and support calculation of demographic indicators. Sites without multi-year data are generally newer or lacked records for earlier years.

### Data ecosystem

All sites reported use of electronic data capture tools such as ODK, SurveyCTO, and REDCap. Nevertheless, one-third of sites still employed parallel paper-based tools for specific modules, which introduces potential for transcription errors and delays.

Integration of complementary data sources such as facility records and environmental datasets was inconsistent. Only a minority of sites routinely used cloud platforms, with most relying on on-site servers and institutional archives. With respect to governance and security, anonymization for external sharing was universal and access-control policies were common. However, formal metadata standards and interoperability with national systems such as DHIS2 were limited. Overall, the data ecosystem showed strengths in adoption of electronic data collection but weaknesses in interoperability, metadata, and cloud-enabled scalability.

#### Data collection

Most HDSS sites (11 of 12) conduct core population and health surveillance activities, including censuses, household surveys, and vital event tracking. Many also collect socio-economic or behavioral data, but far fewer draw on routine health system or environmental sources. Only three sites reported using routine district health data and three reported environmental or geographic data, reflecting variation in integration with national systems. All sites use electronic data collection tools such as CAPI, though five reported supplementing these with paper forms. Alternative data collection methods, such as sensor data or API scraping, are rarely used. Field enumerators remain the backbone of data collection in almost all sites, often supported by community health workers or household representatives. Internal research teams contribute to data processing, while routine health facility staff play only a minor role. Overall, data collection remains centered on trained field teams and direct population surveillance, consistent with HDSS practice.

#### Data handling and quality

Most HDSS sites (11/12) use standard statistical software (Stata, R, SPSS) for cleaning and analysis, with many also relying on relational databases. About half still use spreadsheets while only a few sites employ custom scripts or specialized cleaning tools, reflecting limited integration or automation capacity. Data validation practices differ considerably. Six sites conduct formal checks, ranging from daily to quarterly, while others validate data only annually. Validation commonly occurs both at data entry and during database loading, and most sites that validate data also document their procedures. Although staff report generally high awareness of data quality, common challenges include limited staffing and training, inconsistent data formats, recall bias, and occasional technical or system-related issues.

#### Data management infrastructure and governance

HDSS sites show wide variation in IT capacity, with storage ranging from under 1 TB to over 50 TB. Most sites rely on institution-funded hardware and local servers, while cloud use remains limited. Only four sites reported using platforms such as Azure or Google Cloud. All sites implement core security measures, including data anonymization and backups, and most have access-control policies, encryption, and multi-factor authentication in place. Governance practices are more uneven. While many sites use data-use agreements, IRB oversight, and policies on data ownership, only about half have formal governance frameworks or conduct regular training. Dedicated governance committees and performance monitoring are reported at only a few sites (4/12). Overall, security practices are strong, but governance structures and systematic oversight vary considerably across sites.

#### Synthetic data

Use of synthetic data across HDSS sites is minimal. Only five of the 12 sites reported any need for synthetic datasets, and among them, most rely on simple statistical modeling approaches. Only one site reported needing synthetic data and only one site (The Gambia) identified synthetic data generation as part of its anonymization process. Overall, synthetic data remain rarely used, with most sites continuing to depend on real, anonymized datasets rather than advanced data-synthesis techniques.

#### Data access and sharing

All HDSS sites share data internally, and nearly all (11/12) also share data externally with government ministries or research partners, typically under formal agreements such as Data Transfer Agreements or MOUs. Most sites have data-use and data-ownership policies, though fully open public access is rare. Institutional repositories generally operate under restricted or confidential access controls. All sites anonymize data prior to external release, using methods such as suppression, encryption, subsampling, or hashing. Access decisions are usually overseen by principal investigators or institutional data/security teams. Overall, data sharing is widespread but tightly regulated through agreements and privacy safeguards rather than open public release.

### Data capability maturity

The data capability maturity assessment revealed substantial variability across HDSS sites and four domains (Business Processes, Data Activities, People and Culture, and Technology). Scores ranged from approximately 1.6 to 3.9 on a five-point maturity scale. Only one site (AL-SEHA) approached a proactive maturity profile, while most sites clustered in the stable-to-reactive spectrum. People and Culture emerged as the weakest domain, indicating gaps in leadership engagement, institutional data literacy, and a culture of data-driven decision-making. Several HDSS systems still operate with limited leadership engagement in data governance, variable data literacy among staff, and modest institutional support for data-driven processes. A small number of sites, notably AL-SEHA and Ruwenzori, demonstrated an emerging culture that is increasingly oriented toward structured data governance and evidence-based decision-making. Data Activities and Technology displayed wide disparities, reflecting differences in historical investment and staffing. Business Processes tended to be moderately developed but lacked consistent automation and quality assurance mechanisms.

Site-level analysis showed that AL-SEHA HDSS consistently scored highest across the four domains and approached proactive maturity (Figure 1). Basse, Karonga, and Ruwenzori formed a second tier of higher-performing sites. The mid-tier included Quelinmane, Ouagadougou, Hararghe, and Kyamulibwa, which exhibited functional operations but fragmented processes. The lower tier comprised Niakhar, Taabo, Iganga-Mayuge, and Magu, which remained in the reactive range. The composite maturity index, synthesizing the four domain scores, indicated that most sites are stable but not yet proactive, with only a few demonstrating readiness for AI-enabled workflows and predictive analytics. This pattern suggests a network in transition, with a need for differentiated interventions, i.e., foundational infrastructure and governance for lower-maturity sites and advanced automation and analytic training for higher-maturity sites.

**Figure 1.**
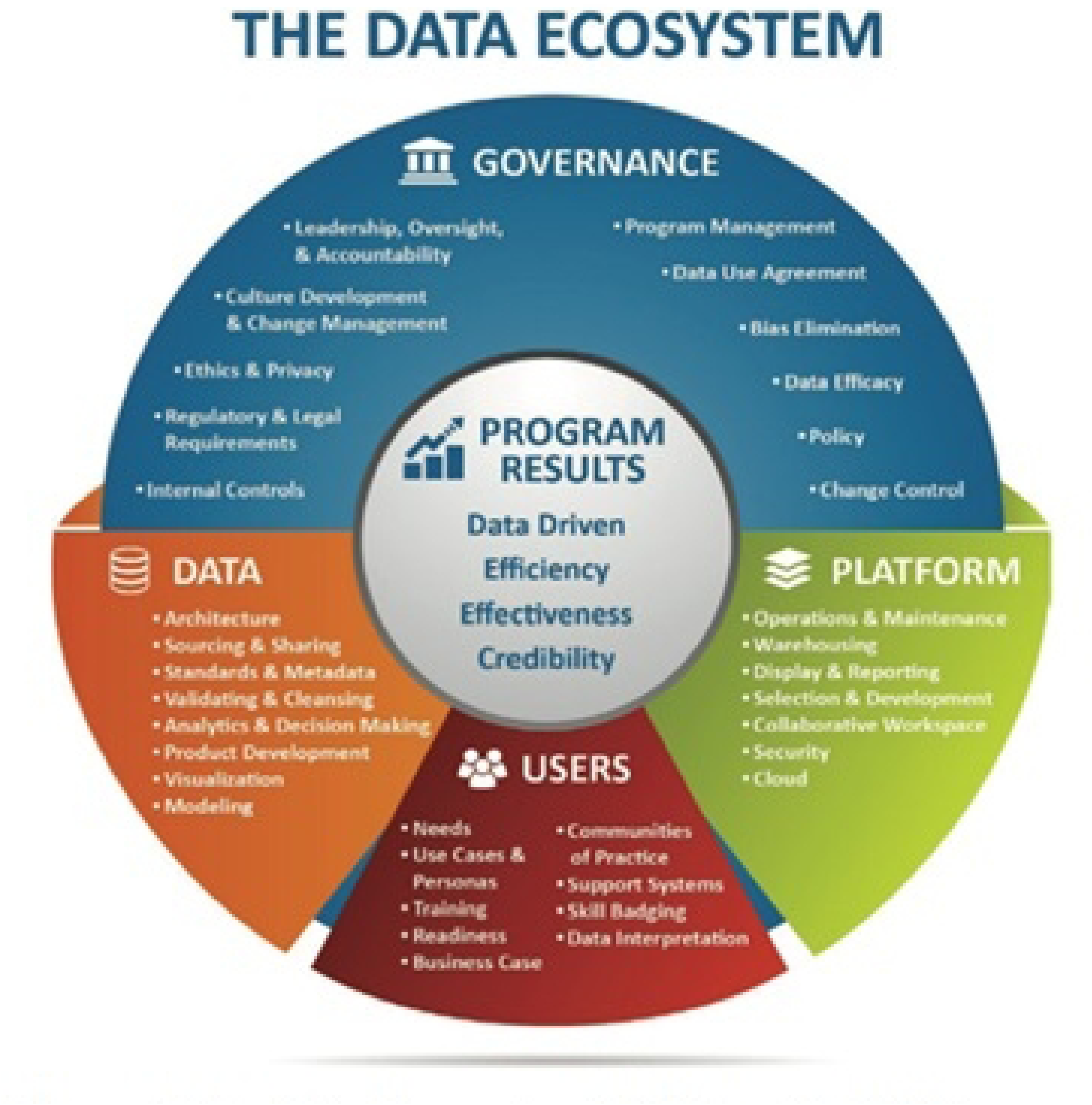
The Data Ecosystem (Malikireddy, 2022)

Data activities exhibited wide variation. Some sites, i.e., Karonga, Basse, Ruwenzori, and AL-SEHA, approached the upper end of the stable or early proactive range, reflecting progress toward structured data workflows, improved metadata documentation, and enhanced quality assurance practices (Figure 1). Others continue to rely on manual processes, inconsistent use of statistical software, and limited implementation of reproducible analytic workflows. Table 1 below summarizes capability maturity scores by site and domain. Scores are presented on a five-point scale with interpretive labels: 1 chaotic, 2 reactive, 3 stable, 4 proactive, and 5 predictive.

**Table 1.** Average capability maturity scores across domains by HDSS site.

| **HDSS Site** | **People & Culture** | **Data Activities** | **Business Processes** | **Technology** | **Overall Maturity** |
| --- | --- | --- | --- | --- | --- |
| AL-SEHA | 3.9 | 4.0 | 3.8 | 4.0 | 3.9 |
| Basse | 3.4 | 3.7 | 3.5 | 3.8 | 3.6 |
| Karonga | 3.2 | 3.8 | 3.4 | 3.6 | 3.5 |
| Ruwenzori | 3.6 | 3.9 | 3.3 | 3.5 | 3.6 |
| Quelimane | 2.8 | 3.2 | 2.9 | 3.0 | 3.0 |
| Ouagadougou | 2.9 | 3.1 | 3.0 | 3.1 | 3.0 |
| Hararghe | 3.0 | 3.2 | 3.1 | 3.0 | 3.1 |
| Kyamulibwa | 2.3 | 2.7 | 2.6 | 2.4 | 2.5 |
| Iganga-Mayuge | 2.1 | 2.6 | 2.5 | 2.3 | 2.4 |
| Taabo | 2.2 | 2.6 | 2.4 | 2.2 | 2.4 |
| Niakhar | 2.0 | 2.4 | 2.3 | 2.1 | 2.2 |
| Magu | 1.8 | 2.1 | 2.0 | 1.9 | 2.0 |

Business Processes were moderately developed across the network. Many sites exhibit functional operational procedures but still lack systematic automation or codified protocols that ensure reproducibility and minimize workflow variability. Several systems demonstrated operational stability but had not yet transitioned to proactive improvement processes. Technology readiness displayed the most pronounced disparities. Sites with strong infrastructural investment, such as AL-SEHA, reached nearly predictive levels of technological maturity with cloud-enabled architectures, robust security layers, and scalable data storage. In contrast, lower-scoring sites such as Niakhar, Taabo, and some of the smaller HDSS platforms demonstrated limited IT infrastructure, minimal automation, and constrained data storage capacity.

### Training needs

Training needs emerged as one of the most significant capability gaps across the HDSS network. The workforce was operationally experienced but unevenly equipped for emerging data science and infrastructure demands. Domains with the highest training needs included technology and infrastructure, data governance, data and analytics, and development and innovation. All sites reported needing at least intermediate training in most domains, with many indicating requirements for advanced or expert-level capacity building in areas such as cloud architecture, automated ETL pipelines, advanced anonymization approaches, and machine learning (Figure 2). A tiered, competency-based training strategy is therefore recommended, combining foundational modules for lower-maturity sites and advanced technical workshops for higher-performing HDSS. Also, training delivery should be bilingual and utilize blended learning approaches to maximize reach and sustainability.

**Figure 2.**
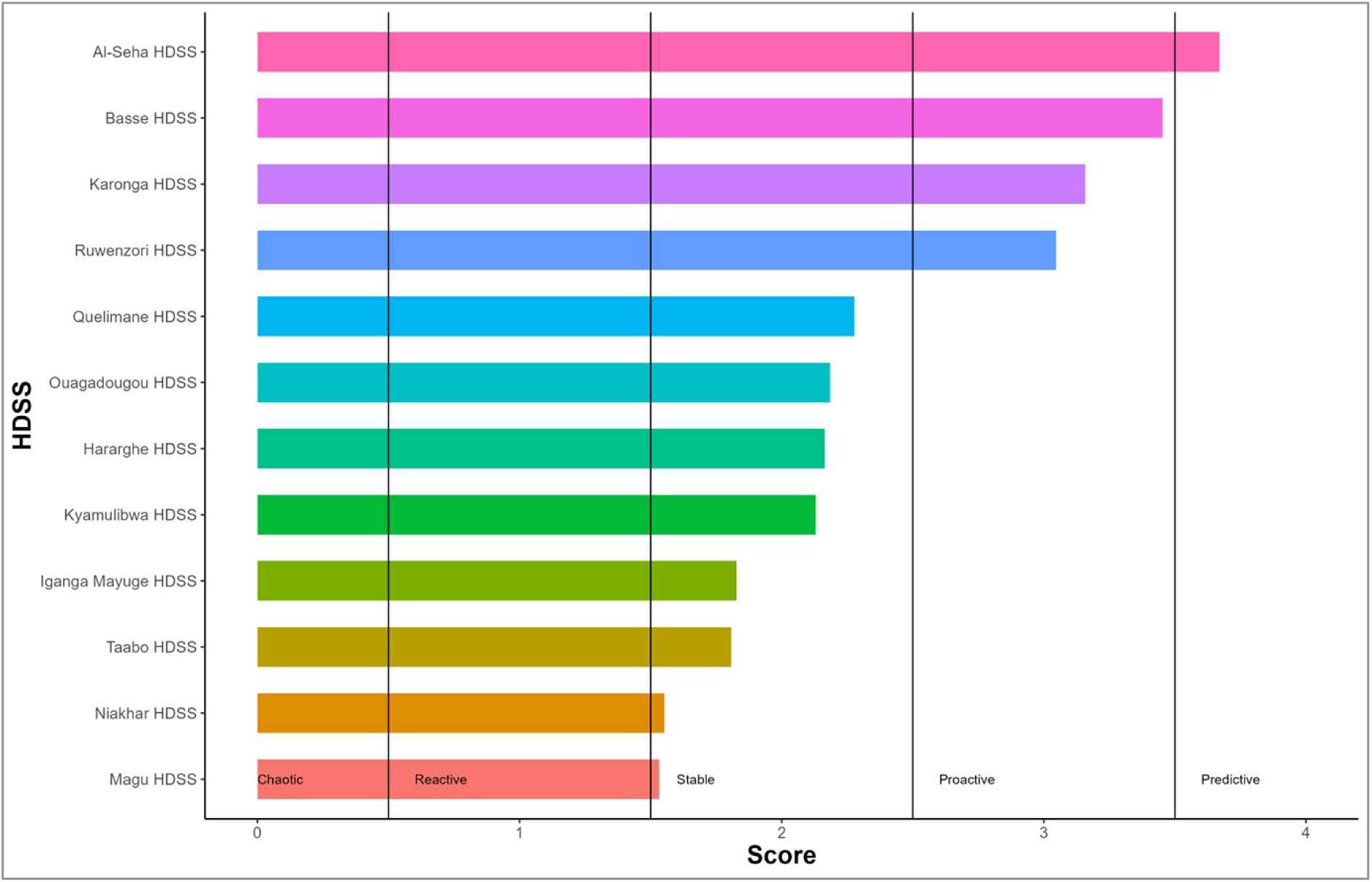
Capability maturity across the HDSS sites.

#### Training demand and competency gaps across HDSS sites

The training needs assessment reveals consistently high demand for capacity-building across all seven competency domains, i.e., 1) Research and Methodology, 2) Technology and Infrastructure, 3) Data and Analytics, 4) Data Governance, 5) Development and Innovation, 6) Project Management, and 7) Visualization and Reporting. Across the network, most sites reported requiring at least intermediate-level training, with many indicating a need for advanced or expert-level support. Almost no domain registered “no training needed” responses, underscoring that skills gaps are widespread and systemic rather than isolated to a few sites. The most significant needs cluster around Technology and Infrastructure, Development and Innovation, Data Governance, and Data and Analytics. It’s important to note that these needs also correspond to the lowest-scoring maturity dimensions identified in the earlier ecosystem assessment.

Patterns in capability and maturity provide further context to the training needs. Maturity scores for Platforms and for User, Culture, and Site Needs tend to be higher than those for Data and Data Governance, pointing to pronounced structural and technological weaknesses. In the Data domain, scores generally fall between 2.5 and 4.0 on a five-point scale, with site-level variation reflecting uneven exposure to training and differing levels of investment in data engineering and management. Governance maturity is similarly varied but with lower overall ceilings, indicating deficits in regulatory compliance, metadata standards, anonymization techniques, and broader institutional governance frameworks. These results suggest that workforce development in data governance is essential for achieving interoperable, ethical, and high-quality data practices across the network.

**Figure 3.**
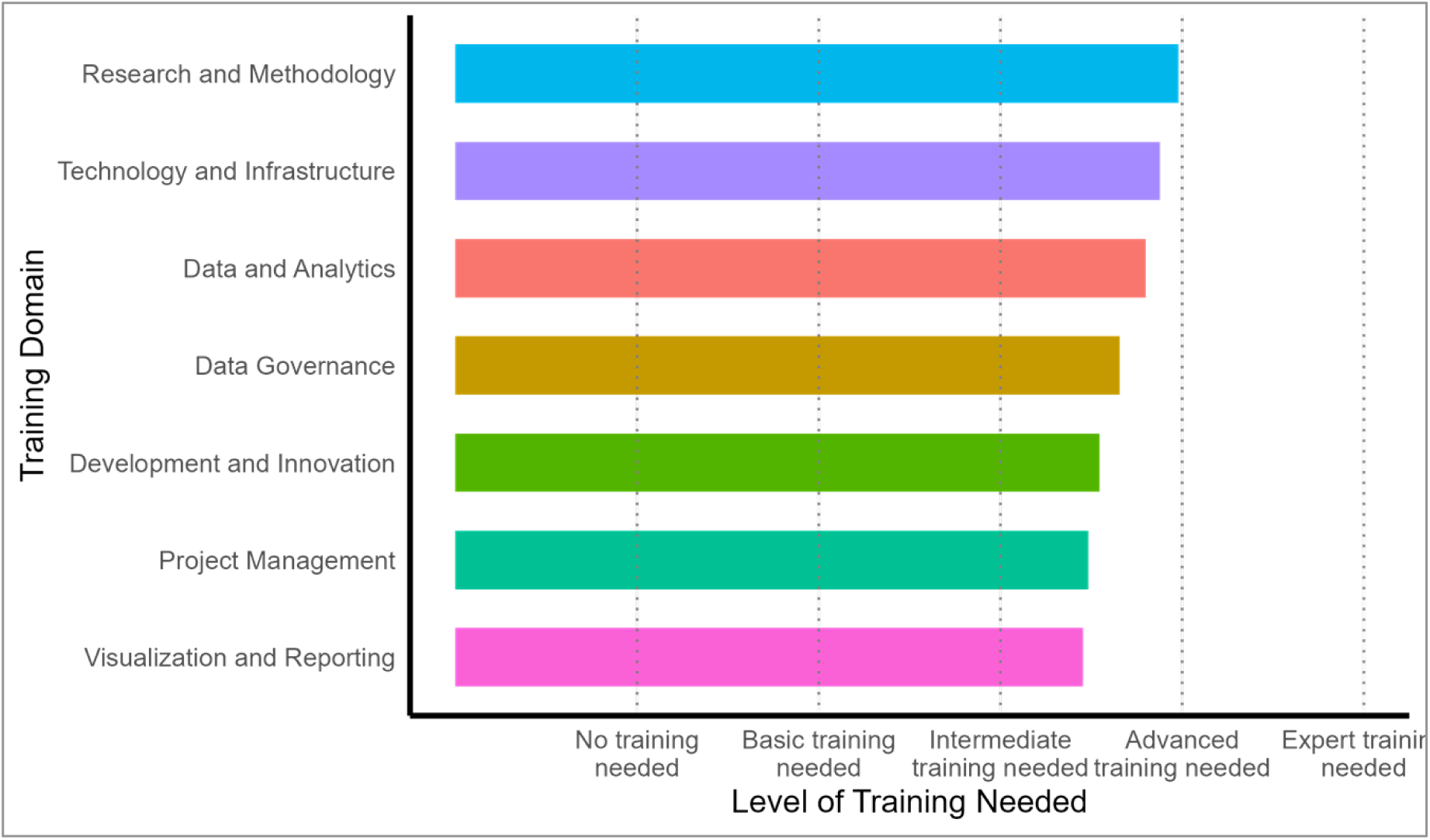
Overall average training needs.

At a more granular level, domain-specific scores show moderate competence but insufficient depth in several technical areas. Training needs in Data and Analytics remain substantial, particularly in statistical programming, data wrangling, modeling, machine learning, and advanced demographic estimation. Some sites report relatively stronger analytics capacity, but all remain below upper proficiency thresholds. Governance competencies also require significant strengthening, especially in metadata design, governance compliance, and advanced privacy-preserving methods. Technology and Infrastructure presents some of the widest disparities across sites, with gaps in cloud architecture, server administration, cybersecurity, and platform optimization. Development and Innovation capacities are constrained by limited experience with automated ETL/ELT pipelines, AI-based harmonization, custom application development, and integration of multiple data streams, including administrative and environmental sources.

Research and Methodology, while a comparatively stronger domain, still shows gaps in longitudinal modeling, mixed-methods integration, causal inference, and the design of complex population-based studies. Project management skills are generally at an intermediate level but would benefit from structured training in multi-site coordination, agile project management, and planning for complex data-collection cycles. Visualization and reporting capacities also lag behind contemporary standards, with several sites requiring training in interactive dashboards, automated reporting pipelines, and tools such as ggplot2, Shiny, PowerBI, and Tableau.

## Discussion

This assessment demonstrates that HDSS platforms across Africa have achieved substantial operational stability but remain at varying stages of maturity with respect to data ecosystems. Key systemic limitations include uneven leadership engagement, incomplete metadata documentation, limited automation, inconsistent governance monitoring, and constrained analytic capacity. These findings align with broader evidence that population data systems in LMICs often face structural barriers to modernization. Addressing these challenges requires a multi-pronged approach that integrates infrastructure investment, standardized metadata and interoperability frameworks, workforce development, and the adoption of automated and AI-enabled data workflows. INSPIRE 2.0 can play a central role by implementing targeted technical assistance, developing bilingual training curricula, and piloting automated harmonization tools at higher-maturity sites before scaling to lower-maturity contexts. Such an approach balances feasibility with ambition and promotes sustained capability building across the network.

### Site characteristics

The HDSS network comprises well-established surveillance systems that consistently generate essential demographic and operational data. Although sites vary in size and context, they share many strengths, including multi-year population monitoring, diverse funding and partnerships, annual census rounds, and substantial investments in data management staff and infrastructure. The 2025 data highlight both current capacities and recent growth, particularly in population coverage and the sophistication of data systems. Sites with more mature data units were able to provide richer, more granular longitudinal indicators, whereas newer or smaller sites showed gaps that signal opportunities for targeted capacity building. Strengthening data management structures across all sites will help ensure comparable detail and quality in future surveillance efforts. The network’s expanding populations, established surveillance practices, and collaborative institutional support position these HDSS sites as critical contributors to longitudinal health and demographic research and to addressing emerging public health challenges.

### Data Ecosystem

The data ecosystem assessment shows that HDSS sites have strong foundations in traditional longitudinal surveillance and have widely adopted digital data collection tools. Most sites practice internal data sharing and maintain anonymization procedures and governance agreements, reflecting strong baseline data stewardship. However, maturity levels vary substantially in more advanced areas of data management. Key gaps include the absence of formal metadata standards, limited interoperability frameworks, and inconsistent governance metrics. Regional patterns are evident as West African sites tend to incorporate more socio-economic or environmental data, while East African sites interact more closely with national health information systems. But overall the heterogeneity across sites underscores the need for harmonization. Improving metadata practices, interoperability, and governance capacity will be essential for enabling sites to fully leverage their longitudinal datasets while ensuring privacy, compliance, and cross-site comparability.

### Data maturity

The maturity assessment shows that while HDSS sites have achieved operational stability, they have not yet developed the technical depth, analytical capacity, or automation needed to fully align with modern population data science systems. Persistent gaps in leadership culture, metadata quality, automation, and advanced analytics continue to limit progress toward higher maturity. These findings indicate that INSPIRE 2.0 should pursue a differentiated capacity strengthening strategy, i.e., providing foundational support to lower-maturity sites while offering more advanced, specialized interventions to higher-performing ones. Tailoring capacity-building efforts in this way will promote sustainable growth, strengthen harmonization readiness, and lay the groundwork for AI-enabled data activities across the network.

### Training needs

While HDSS sites possess strong foundations in field operations and basic data handling, significant skill gaps remain in areas essential for modern, interoperable population data systems. These gaps constrain progress toward higher maturity functions such as cloud-based infrastructure, automated data engineering, harmonized workflows, and AI-enabled analytics. To address this, INSPIRE 2.0 will need to implement a tiered, competency-based training strategy that supports foundational upskilling for lower-maturity sites while providing advanced technical, governance, and architectural training for more mature sites. The bilingual nature of the network further underscores the need for flexible, context-specific training approaches that accommodate diverse institutional capacities.

### Study Limitations

This assessment has several limitations that should be considered when interpreting the findings. First, the analysis relied primarily on self-reported data from HDSS site leads and technical staff, which may be subject to reporting bias or differences in how capability levels were interpreted across sites. Second, although the assessment covered 12 HDSS sites, the maturity scores and training needs may not fully represent the heterogeneity of all African HDSS platforms, particularly newer or smaller sites not included in this assessment. Third, quantitative data on infrastructure, staffing, and data systems were not independently verified through direct audits, which may have led to under- or overestimation of capability levels. Fourth, the absence of qualitative data limits deeper exploration of contextual factors, such as organizational culture, funding variability, or historical system challenges, which all shape data capability maturity. Finally, because the assessment reflects a cross-sectional snapshot, it cannot capture temporal changes or ongoing improvements occurring after data collection. Despite these limitations, the study provides an important comparative baseline for strengthening HDSS capacity in Africa.

## Conclusion and Recommendations

This multi-country assessment reveals that the assessed HDSS are not well positioned for the evolving demands of modern population data science. Across Africa, HDSS platforms have built strong operational foundations through stable surveillance cohorts, longstanding field presence, and deep experience generating longitudinal population data. But the assessment also makes clear that structural and systemic disparities persist, shaping how effectively sites can transition toward more integrated, automated, and analytically advanced data ecosystems.

Patterns across domains showed that the maturity of a site’s data ecosystem was closely related to its organizational characteristics, workforce capacity, and degree of alignment with national information systems. Larger or more established HDSS sites tended to exhibit more advanced governance frameworks, more complete metadata practices, and stronger analytic pipelines, whereas newer or smaller platforms often struggled with infrastructure limitations, insufficient staffing depth, and inconsistent documentation standards. These variations point to the need for strengthening technical and leadership capacity in tandem, as sites with higher engagement from institutional leadership were more likely to demonstrate readiness for modernization and cross-site interoperability.

The assessment also identified shared bottlenecks such as incomplete metadata standards, limited automation, and inconsistent governance monitoring, all of which hinder harmonization and restrict the extent to which HDSS data can be easily integrated across sites or leveraged for real-time insights. At the same time, regional differences in data sources, national system linkages, and the breadth of social or environmental indicators collected suggest that modernization efforts must be sensitive to HDSS contexts. Efforts to strengthen interoperability and standardization will need to balance uniformity with sufficient flexibility to accommodate local priorities and data environments.

Close alignment between maturity gaps and training needs further underscores the central role of workforce development in enabling ecosystem transformation. Skills shortages in automation, cloud systems, metadata engineering, and advanced analytics map directly onto the domains where capability maturity was lowest. These correlations point toward the importance of structured, multi-level trainings that support both foundational upskilling and more specialized technical training for higher-performing sites. Discrepancies between perceived and demonstrated capability also suggest that capacity-strengthening programs should incorporate opportunities for self-assessment and reflection to help HDSS teams recalibrate internal expectations and identify areas where external technical support may be most impactful.

Looking ahead, the findings highlight several strategic directions for accelerating progress across for HDSS in Africa. First, modernizing metadata standards and strengthening governance mechanisms should be prioritized as basic steps towards harmonized analytic workflows and secure data exchange. Second, investments in infrastructure, including cloud-enabled architectures, secure storage solutions, and scalable computing environments, will be necessary to support automation and more advanced analytical methods. Third, the introduction of AI-assisted data harmonization, automated quality checks, and operation workflows should begin with higher-maturity sites that are ready to pilot such innovations, with lessons learned subsequently transferred to lower-maturity sites. Fourth, the implementation of bilingual, context-sensitive training programs will be essential to support workforce readiness and ensure equitable access to capacity-building opportunities across Francophone and Anglophone regions. Finally, building communities of practice that encourage cross-site collaboration, peer learning, and shared problem-solving will help sustain momentum and foster a culture of continuous improvement.

In conclusion, HDSS platforms across Africa are at a pivotal point. With well-coordinated investments in governance, metadata, infrastructure, and workforce development, implemented through a differentiated, maturity-aligned strategy, these systems can evolve from traditional surveillance platforms into a robust, harmonized continental data infrastructure. INSPIRE 2.0 is uniquely positioned to catalyze this transformation by providing strategic direction, targeted technical support, and a collaborative framework that promotes innovation and interoperability. By advancing these priorities, the HDSS can strengthen their role as a cornerstone of population research and policy decision-making in Africa.

## Ethical considerations

Ethical approval was obtained from AMREF-IERC while all participants provided informed consent. Data were handled according to HDSS ethical frameworks.

## Data availability

The data underlying this article are available from the corresponding author on reasonable request and subject to HDSS data governance policies and data sharing agreements.

## Author contributions

Conceptualization: Agnes Kiragga. Methodology: Samuel Iddi, Ivan Busulwa, Rachel Odhiambo, Daniel Maina. Data curation: Rachel Odhiambo, Henry Odero. Formal analysis: Rachel Odhiambo. Writing (original draft): Ivan Busulwa. Writing (review & editing): Samuel Iddi. Supervision: Agnes Kiragga.

